# Biomarkers of the Microbiome-Skin-Brain Axis in Stress and Depression: Fingerprinting of Highly Volatile Compounds in Axillary Sweat via Gas Chromatography-Ion Mobility Spectrometry

**DOI:** 10.1101/2025.05.06.25327121

**Authors:** Nuttanee Tungkijanansin, Chadin Kulsing, Chavit Tunvirachaisakul, Sira Sriswasdi, Stephen J. Kerr, Jidapa Hanvoravongchai, Napatsorn Thewaran, Patthrarawalai Sirinara, Michael Maes

## Abstract

**Background:** Difficulty in the diagnosis of high stress and depression has been recognized conventionally depending on the observation of patient symptoms and psychiatrist diagnosis. These approaches are time-consuming and cannot respond to the excessive demands for large-scale tests with the increasing populations worldwide. This study thus developed an alternative approach to perform fast stress screening, which is based on fingerprinting of highly volatile compounds in axillary sweat.

**Methods:** Sweat samples were collected from 227 firefighters, comprising 65 with high stress, 14 with depression, and 148 healthy volunteers. High stress and depression were determined using the standardized Thai versions of the Perceived Stress Scale (PSS-10) and the Beck Depression Inventory II (BDI-II), in conjunction with psychiatric interviews. The samples were collected by placing cotton rods under the axillaries, then analyzed using gas chromatography– ion mobility spectrometry (GC-IMS). The potential marker peaks were selected based on accuracy data. Principal component analysis (PCA) and logistic regression with machine learning were also performed to select significant composite markers. MVOC 3.0, Amibase and Metaboanalyst 6.0 databases were applied to predict the possible metabolomic pathways.

**Results:** Analysis against genuine standard compound injections identified acetonitrile, ammonia, diethyl ether, formaldehyde, and octane as potential biomarkers for both high stress and depression, with butane, dimethylamine and pentane additionally observed for high stress. Receiver operating characteristic (ROC) curves demonstrated accuracies of 81.3% for stress screening and 82.8% for depression screening.

**Conclusion:** The biomarkers delineated here indicate the participation of particular metabolic pathways and commensal skin bacteria in the stress response.

## Background

Existing epidemiology evidence has reported that firefighters have higher rates of mental health problems than the overall population.^1–9^ High rates of potentially traumatic exposure, high-stress situations, and sleep disruption harm the mental health of firefighters.^10–14^ In Korea, recent research pointed out that the risk of suicidal ideation associated with occupational stress and depressive symptoms was as high as 1.066% in firefighters.^5^ A study in Japan demonstrated that 22.3% of firefighters exhibited depressive symptoms related to their job stress and high workload.^15^ Similarly, a high prevalence of suicidal ideation and attempts (46.8% and 15.5%, respectively) related to occupational stress was found among U.S. firefighters. ^4,6^ At least 19% of UK firefighters self-reported having at least one mental health issue as a result of high stress. ^2^ Psychological evaluation for public safety occupations with high mental illness rates, such as firefighters, demands easy-to-access and reliable mass screening.

To date, there is little knowledge regarding an objective, unbiased, highly sensitive and specific screening tool. Emerging evidence from interdisciplinary research indicates the existence of communication axes between the brain-gut and skin-brain axis.^16–18^ Several approaches have been established for high stress and depression diagnoses such as blood levels or wearable devices.^19–21^ Alternatively, a non-invasive method analyzing volatile compound markers for stress has been reported. Gas chromatography-mass spectrometry (GC-MS) and statistical data analysis have been widely applied for tentative identification of the volatile markers by comparing the experimental MS spectrum and retention index data with the libraries. Among different body fluids, sweat analysis is attractive in the aspects of non-invasive, safe sample preparation protocol (such as negligible amounts of Corona virus),^22^ extractable onto substances facilitating automated analysis. Volatile compounds could be originated from the apocrine glands. ^23^ Alcohols and aldehydes are metabolites that are produced by cutaneous microorganisms through the interaction with non-volatile compounds from the sebaceous gland. ^24–26^ For example, triglycerides could be hydrolyzed by lipases produced from anaerobic bacteria leading to the long-chain acids, and these acids could be further converted by aerobic bacteria into the shorter chain saturated and unsaturated acids and the volatile aldehydes and alcohols. ^26^ Because it is reasonable to anticipate that the microbiome compositions of populations with elevated stress or depression will be distributed differently in their sweat, their sweat volatile compound profiles may also differ. ^27,28^ This suggests it is possible to discover sweat volatile compound markers for stress or depression. For example, increased amounts of n-decanoic acid were reported as a possible stress marker. ^29,30^ Martin et al. recruited fifteen young adult volunteers (19–26 years) for two randomized crossover interventions. They employed a “timed audio serial addition task” intervention that required participants to perform a series of quick mental arithmetic computations that induced stress, whereas the “neutral” intervention had participants listen to peaceful music. Each intervention used skin patches to collect skin-VOC samples, which were then frozen at -80 °C and analyzed using GC-MS. The VOC *n*-decanoic acid was found to be upregulated. VOC profiles in skin appear to vary in response to stress due to increased excretion, higher bacterial activity, or possibly increased oxidative pathways. ^29^

Alternatively, n-decanoic acid may also be produced by microbiomes such as *Streptomyces* and *Filamentosus* (www.amibase.org). Increased benzoic acid in sweat as well as decreased xylene and 3-carene contents in sweat may result from psychological stressors. ^29,30^ These compounds are not generated during human metabolisms, suggesting that they are formed by other pathways. For example, *Bacteroides, Enterobacteriaceae, Veillonella, Streptococcus* and *Clostridium* (www.amibase.org) could produce benzoic acid. The change in amounts of “*Esherishia. coli*” and “*Bacteroides, Enterobacteriaceae, Veillonella, Streptococcus, Clostridium*”, respectively, could also be responsible for the lower amounts of xylene and 3-carene (www.amibase.org). The discovery of highly volatile markers in sweat for depression screening is still a challenging task due to the limited capability of the conventional gas chromatography-mass spectrometry (GC-MS) in analysis of samples with coeluting components ^31,32^ especially for the untargeted identification of highly volatile compounds.

GC-ion mobility spectrometry (GC-IMS) can be effectively applied to analyze highly volatile compounds based on the two-dimensional separation capability of GC (based on boiling point/interaction with the stationary phase) and IMS (based on several parameters such as ion charges and collisional cross sections between the ions and the drift gas molecules). This approach results in different retention times (*t*_R_) and drift times (*t*_dr_) of each compound. For example, GC-IMS applications include identification of ethanal, octanal, acetone, butanone, and methanol as the potential markers in breath for screening of SARS-CoV-2 infection, ^33^ as well as other screening tests for pulmonary diseases, infections and toxins. ^34^ GC-MS analysis was also applied to investigate anxiety sweat with the related compounds of pentadecane, 4,6-dimethyldodecane, dodecanal, 1-dodecanol, hexadecane and tetradecanal. ^35^ In the latter study, stress and anxiety were induced by the Trier Social Stress Test (TSST).

In the current study, we measured volatile compounds using GC-IMS in axillary sweat of firefighters with and without stress or depression. Based on these results, we selected a panel of the best performing biomarkers of stress and depression. These biomarkers were selected using machine learning methods and a comparison of the *t*_R_ and *t*_dr_ values with the authentic standards. The screening capability was then investigated via the construction of receiver operating characteristic (ROC) curves.

## Methods

The study protocol, including administration of the questionnaire, collection of axillary sweat samples, and psychiatric diagnoses, was approved by the Institutional Review Board of the Faculty of Medicine, Chulalongkorn University (IRB No. 931/64). Prior to participation, all individuals received information regarding the study’s objectives and design, and provided written informed consent. This research was conducted in accordance with the Helsinki Declaration.

### 1. Investigated population and measurement tools

The study population consisted of currently employed professional firefighters from 48 fire stations listed in the Bangkok fire and rescue department registry, which is a part of the Bangkok Metropolitan Administration (BMA) (n = 227). All firefighters 18 years of age or older who tested negative for COVID-19 and provided written informed consent were eligible for inclusion in the study. Firefighters were excluded if they had an annual physical ability test score below 70%, or if they were classified as administrative personnel or volunteer firefighters. A station-based or cluster sampling approach was used, with a robust participation rate 60–70% in each station.

Stress was assessed using the standardized Thai version of the Perceived Stress Scale (PSS-10) questionnaire. This self-rated questionnaire consists of 10 items that score the degree of perceived stress of life events. Scoring of the answers is based on a 0 to 4 scale for each item. The PSS-10 has good internal consistency (Cronbach’s alpha =0.85) and a content validity of 0.82 for measuring the degree of perceived stress in a Thai population.^36^ Firefighters with a score of less than or equal to 31 were identified as the no-stress group, whereas those with scores 32-40 were classified as having high stress.^36^

The Thai version of Beck Depression Inventory version II (BDI-II) was utilized to assess the severity of depression of firefighters during the past two weeks. The 21-item self-report questionnaire is weighted on a 0–3 interval scale from 0 (no symptom) to 3 (most severe), with higher scores indicating greater feelings of depression. The BDI-II scores correlate significantly with the Hamilton Depression Rating Scale (HDRS) scores (*r* = 0.72, *p* < 0.0001) ^35^. Depression severity is assessed as follows: 1-9 no depression, 10-15 mild depression, 16-19 mild-to-moderate depression, 20-29 moderate-to-severe depression, and 30+ severe depression. The BDI-II demonstrates good internal consistency (Cronbach’s alpha =0.89).^37^ Previous studies showed that the PSS-10 had positive correlation with depression.^38–40^ These findings are in line with the report of Cohen et al. that there is some overlap between depressive scales and the PSS-10, since stress may be a symptom of depression.^41^

Participants were initially screened with a questionnaire that categorized them as having either high or normal stress and either depression or no depression. Two blinded investigators compared all data coded, blank or multiple responses to the original response forms and corrected any possible errors. Individuals who screened positive then underwent a confirmatory diagnostic interview with a psychiatrist using DSM-5 criteria.

### 2. Sweat sample collection

For each volunteer, a sweat sample was collected using two cotton rods (0.8 x 2.5 cm) held under the left and right axillarys for 15 min (**Figure 1**). The two cottons containing the axillary sweat were then inserted into a 20-mL screw thread headspace glass vial closed with an aluminum cap with sealed PTFE/silicone screw septa (National Scientific, Rockwood, TN, USA) held by trained staffs. The samples were kept in the refrigerator. The vials were equilibrated at room temperature (30 °C) for at least 15 min before GC-IMS analysis.

### 3. HS GC-IMS analysis

The sample HS were injected into the GC injection port at 70 °C under splitless mode. The volatile components were separated on a FS-SE-54-CB-1 capillary column (15 m x 0.25 mm inner diameter, 0.25 µm film thickness; CS-Chromatographie Service GmbH (Langerwehe, Germany). Ultra-high purify nitrogen gas (99.999% purity) was used as the carrier. The carrier gas flow rate was programmed as follows: 2 mL min-1 for 5 min, increased to 10 mL min-1 within 10 min and then to 20 mL min-1 within 25 min. The GC oven temperature was isothermal at 40 °C during the analytical separation. The oven temperature was then increased to 80°C in order to clean the capillary prior to the next analysis. The drift tube length was 9.8 cm and operated at 45 °C under a 150 mL/ min nitrogen flow. A constant electric field strength of 500 V/cm was applied throughout the analysis. Blank cotton materials were also analyzed under the same experimental condition in order to differentiate the background signals from the sweat components.

### 4. Data analysis

Descriptive statistics were used to summarize demographic, clinical, and volatile organic compound data. Group comparisons were conducted using Student’s t-test for continuous variables and Fisher’s exact test for categorical variables. All analyses were performed using Stata, version 18.0 (StataCorp LLC, USA). A p-value of <0.05 was considered statistically significant. Data presentation, analysis and peak integration were performed by using the LAV software. Significant features (peak positions in contour plots) were selected in order to cover all the space containing peaks of all the samples. Volumes of these features were then measured and used for statistical and machine learning analyses below.

Predictive modeling for high stress and depression outcomes were performed using logistic regression and random forest consisting of 100 decision trees. Recursive feature elimination technique was performed to narrow down the list of informative VOC and clinical variables by starting with the full input data and then iteratively dropping the least important variables from the model. Hyperparameter optimization was performed using grid search on a 3-fold cross-validation strategy. For logistic regression, the regularization strength (from 10^-3^ to 10^5^), penalty function (L1 or L2), and class weight (no weight or weighted by frequency) were tuned. For random forest, the maximum depth (from 2 to 25), the number of input features to explore at each step (from 30% to 70% of the total), the minimum number of samples for creating a split (from 5 to 20 samples), and class weight (no weight or weighted by frequency) were tuned. The best model was selected based on the area under the precision-recall curve (AUPRC) which focuses on the ability to identify rare high stress and depression outcomes. Feature importance and Shapley Additive Explanations (SHAP) values were computed to quantify the contribution of each volatile compound and clinical variable to the prediction of high stress and depression outcomes.

Dimensionality reduction techniques, namely Principal Component Analysis (PCA) and Uniform Manifold Approximation and Projection (UMAP), were applied to visualize the VOC dataset. Hierarchical clustering and spectral clustering (a network clustering technique) were then performed to categorize participants based on their VOC profiles. The optimal number of clusters was determined using the highest Silhouette score and Calinski-Harabasz index (CH index). Associations between VOC and clinical variables and the clusters were assessed using Mann-Whitney U test and chi-squared test. Benjamini-Hochberg procedure was performed to correct for multiple testing with a target false discovery rate of 5%. The sets of VOC markers were related with microbiomes using MVOC 3.0 and Amibase databases and human metabolisms using MetaboAnalyst 6.0.

## Results

Of the 2,036 firefighters employed by the Bangkok Metropolitan Administration, 227 (11.1%) were enrolled in the study, representing the entire workforce across all 48 fire stations in Bangkok. All participants were male, with a mean age of 41.3 years (SD 5.7) (**Table 1**). Approximately 33.5% of participants reported at least one coexisting health condition, with hypertension, gastritis or peptic ulcer, and type 2 diabetes being the most common. Among all participants, 2.6% had a diagnosed mental health disorder, and an equal proportion were receiving psychotropic medications. Smoking status was categorized based on questionnaire responses, defining “smokers” as individuals who had smoked at least 100 cigarettes after initiating smoking and “non-smokers” as those who had either never smoked or had smoked fewer than 100 cigarettes in total. ^42^ Lifetime smoking exposure was quantified using the Brinkman Index, calculated as (number of cigarettes per day) × (number of years for which a person smoked). ^43^ In this study, all participants in this study were current non-smokers.

**Table 1.** Characteristics of firefighters.

| Characteristics | Total (N=227) |
| --- | --- |
| Age (mean±SD) — yr | 41.3±5.7 |
| Males — n (%) | 227 (100%) |
| BMI (mean±SD) — kg/m<sup>2</sup> | 25.3±3.8 |
| Smoking— n (%) | 0 (0%) |
| Drinking alcohol — n(%) | 21 (9.3%) |
| Coexisting conditions— n (%) |  |
| At least one coexisting condition | 76 (33.5%) |
| Hypertension | 31 (13.7%) |
| Gastritis or peptic ulcer | 17 (7.5%) |
| Type 2 diabetes | 11 (4.8%) |
| Cardiovascular disease | 2 (0.9%) |
| Stroke | 4 (1.8%) |
| Diagnosed psychiatric disorders | 6 (2.6%) |
| Current psychotropic medication use — n (%) | 6 (2.6%) |

SD - Standard Deviation, IQR - Interquartile Range

The geographic distribution of axillary sweat collection sites across fire stations in Bangkok, Thailand, is shown in **Figure 1**, with numbered markers representing individual fire stations. The inset provides a detailed illustration of the sweat collection process, including sampling, collection duration, and subsequent GC-IMS analysis.

**Figure 1.**
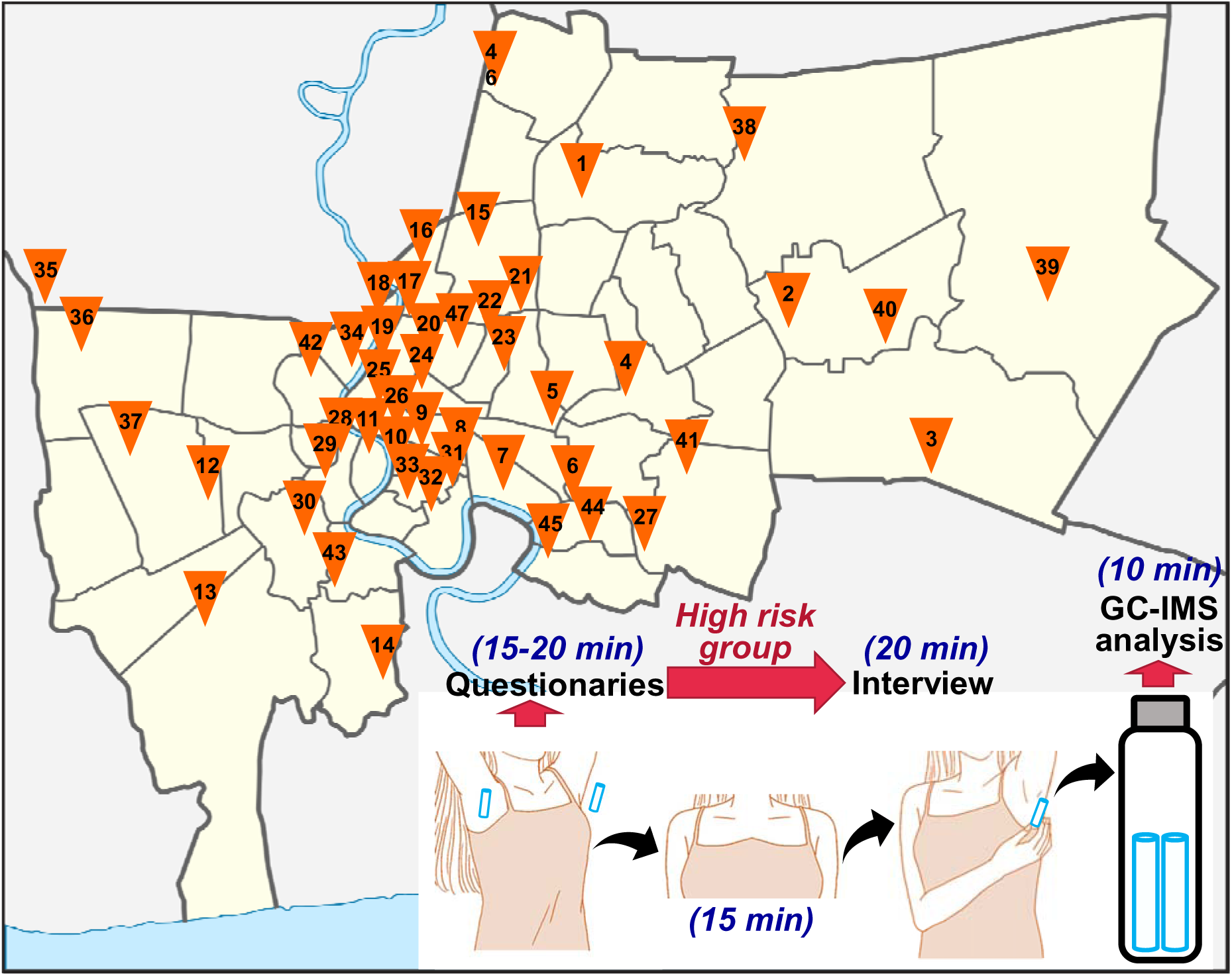
Map of 48 participating fire stations in Bangkok, Thailand, with numbered markers indicating sweat collection sites. The inset illustrates the step-by-step sweat collection process.

### GC-IMS analysis and possible volatile compound markers

All samples were analyzed with GC-IMS, which resulted in *t*_R_ vs. *t*_dr_ vs. intensity contour plots containing peaks (features) with different volumes and positions. This represents fingerprints of different samples according to their different volatile compound profiles as illustrated in **Figure 2**. Samples from participants with high stress or depression showed different fingerprints compared to those without stress or depression.

Such difference is significant and could not be caused by the instrumental error during the repeated analyses, as illustrated by the good repeatability in repeated injections of the same sample within three different days (*n* = 5 within each day). This leads to the intraday and interday relative standard deviations (RSD) of the *t*_R_ of the significantly detected peaks of 0.0-0.1% and 0.0-0.1%, respectively. The corresponding values for *t*_dr_ were 0.0-1.1% and 0.0-0.4%, respectively, and that of the peak volumes were 5.2-13.2% and 4.6-10.0%, respectively.

**Figure 2.**
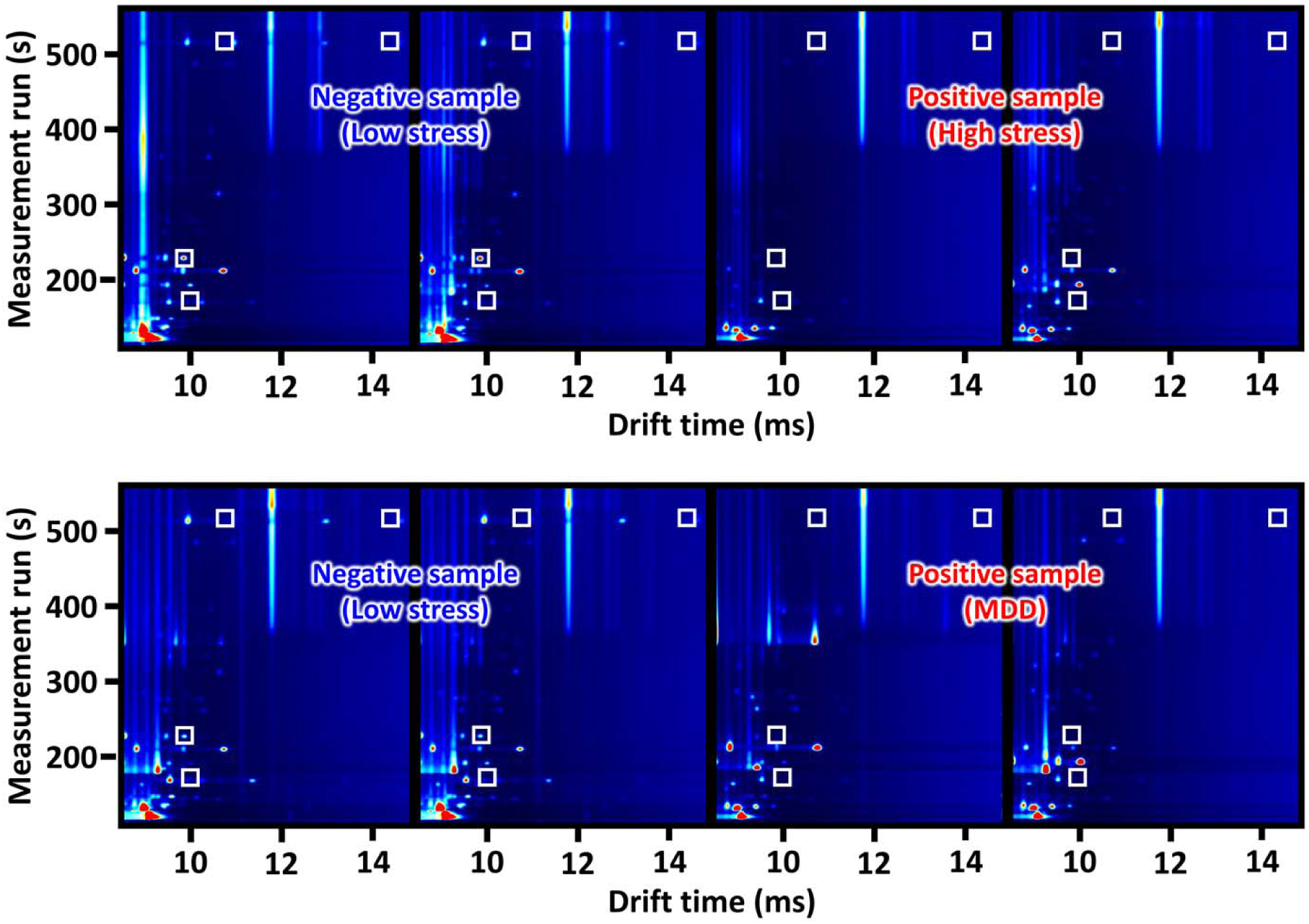
Example GC-IMS contour plots of (A and B) mental stress negative, (C and D) mental stress positive, and (E and F) mental stress and MDD negative axillary sweat samples, with the marker peaks indicated by the squares.

### Stress

There were three firefighters with incomplete PSS-10 scores, therefore 224 were analyzed. There were no significant differences in the distribution of job positions between the high-stress and normal-stress groups. Additionally, the high-stress group’s perceived stress scores are substantially higher (*p* < 0.001), indicating a marked difference in stress perception between the high and normal stress groups. There was a statistically significant difference in regional distribution between the high stress and normal stress groups ( < 0.001). Compared to firefighters with normal stress, those in the high stress group were disproportionately located in Regions 4 and 6. In contrast, Region 5 contained a notably larger proportion of normal stress firefighters (54.9%) than high stress firefighters (22.6%). Other characteristics, such as age and BMI did not show significant differences between the two groups (**Table 2**).

**Table 2.** Characteristics of firefighters with high and normal mental stress.

| Characteristic | high stress<br>(N=62) | normal stress<br>(N=162) | *p*-value<sup>†</sup> |
| --- | --- | --- | --- |
| Age (years) — mean±SD | 42.5±6.1 | 41.3±5.7 | 0.170 |
| Body Mass Index (BMI, kg/m<sup>2</sup>) — mean±SD | 24.9±4.2 | 25.5±3.6 | 0.300 |
| Position — n (%) |  |  |  |
| Division Chief | 0 (0.0%) | 3 (1.8%) | 0.470 |
| Senior firefighter | 1 (1.6%) | 8 (4.9%) |  |
| Operational firefighter | 61 (97.7%) | 152 (93.2%) |  |
| Region — n (%) |  |  |  |
| Region 1 | 5 (8.1%) | 6 (3.7%) | <0.001 |
| Region 2 | 18 (29.0%) | 28 (17.2%) |  |
| Region 3 | 7 (11.3%) | 31 (19.0%) |  |
| Region 4 | 9 (14.5%) | 4 (2.5%) |  |
| Region 5 | 14 (22.6%) | 89 (54.9%) |  |
| Region 6 | 9 (14.5%) | 3 (1.9%) |  |
| Region 7 | 0 (0.0%) | 1 (0.6%) |  |
| Perceived stress scores (PSS-10) —mean±SD | 36.0±2.5 | 24.2±3.7 | <0.001 |

SD - Standard Deviation.
<sup>†</sup> *p*-values for continuous variables were calculated with the use of t-test, and *p*-values for categorical variables were calculated with the use of Fisher's exact test.

### Depression

There were significant differences in gender distribution, region, and Beck Depression Inventory-II (BDI-II) scores between firefighters experiencing depression (BDI-II score ≥ 16) and no-depression (BDI-II score < 16), as demonstrated in **Table 3**. The mean age was significantly higher in the depression group (45.5 years; SD 5.9) compared to the no-depression group (41.4 years; SD 5.7) (*p*=0.015). A significant difference in regional distribution was noted (*p*=0.002). The mean Beck Depression Inventory-II (BDI-II) score was substantially higher among firefighters with depression (22.2; SD 6.9) than among those without depression (1.7; SD 3.1) (p < 0.001). BMI and position of professional firefighters did not show significant differences between the two groups.

**Table 3.** Characteristics of firefighters with and without depression.

| Characteristics | depression<br>(N=14) | no<br>depression<br>(N=213) | *p*-<br>value<sup>†</sup> |
| --- | --- | --- | --- |
| Age (years) — mean±SD | 45.5±5.9 | 41.4±5.7 | 0.015 |
| Body Mass Index (BMI, kg/m<sup>2</sup>) — mean±SD | 25.2±4.1 | 25.3±3.8 | 0.900 |
| Position — n (%) |  |  |  |
| Division chief | 0 (0.0%) | 2 (0.9%) | 0.694 |
| Senior firefighter | 1 (0.4%) | 8 (3.5%) |  |
| Operational firefighter | 13 (5.7%) | 203 (89.4%) |  |
| Region — n (%) |  |  |  |
| Region 1 | 4 (28.6%) | 7 (3.3%) | 0.002 |
| Region 2 | 2 (14.3%) | 45 (21.1%) |  |
| Region 3 | 1 (7.1%) | 37 (14.6%) |  |
| Region 4 | 0 (0.0%) | 13 (6.1%) |  |
| Region 5 | 6 (42.9%) | 100 (46.9%) |  |
| Region 6 | 0 (0.0%) | 12 (5.6%) |  |
| Region 7 | 1 (7.1%) | 0 (0.0%) |  |
| Beck Depression Inventory-II (BDI-II) scores — mean±SD | 22.2±6.9 | 1.7±3.1 | <0.001 |

<sup>†</sup> *p*-values for continuous variables were calculated using t-test, and *p*-values for categorical variables were calculated using Fisher's exact test.

### Association of VOC to stress and depression

There were moderate correlations between VOC and clinical variables (**Figure 3**, Spearman rank correlation coefficients ranging from -0.4027 to 0.2809), including 45 VOC markers with significantly different levels in the sweat of participants with and without diagnosed stress and 10 VOC markers between participants with and without diagnosed depression (adjusted p-value ≤ 0.05, top markers shown in **Supplementary Figure 1**). For stress, area6, area7, and area9 are the best VOC markers with 1.57-1.74 folds lower levels on average in participants without stress. For depression, the best VOC markers are area88 and area44 with 1.46-2.17 folds higher level on average in participants with depression.

**Figure 3.**
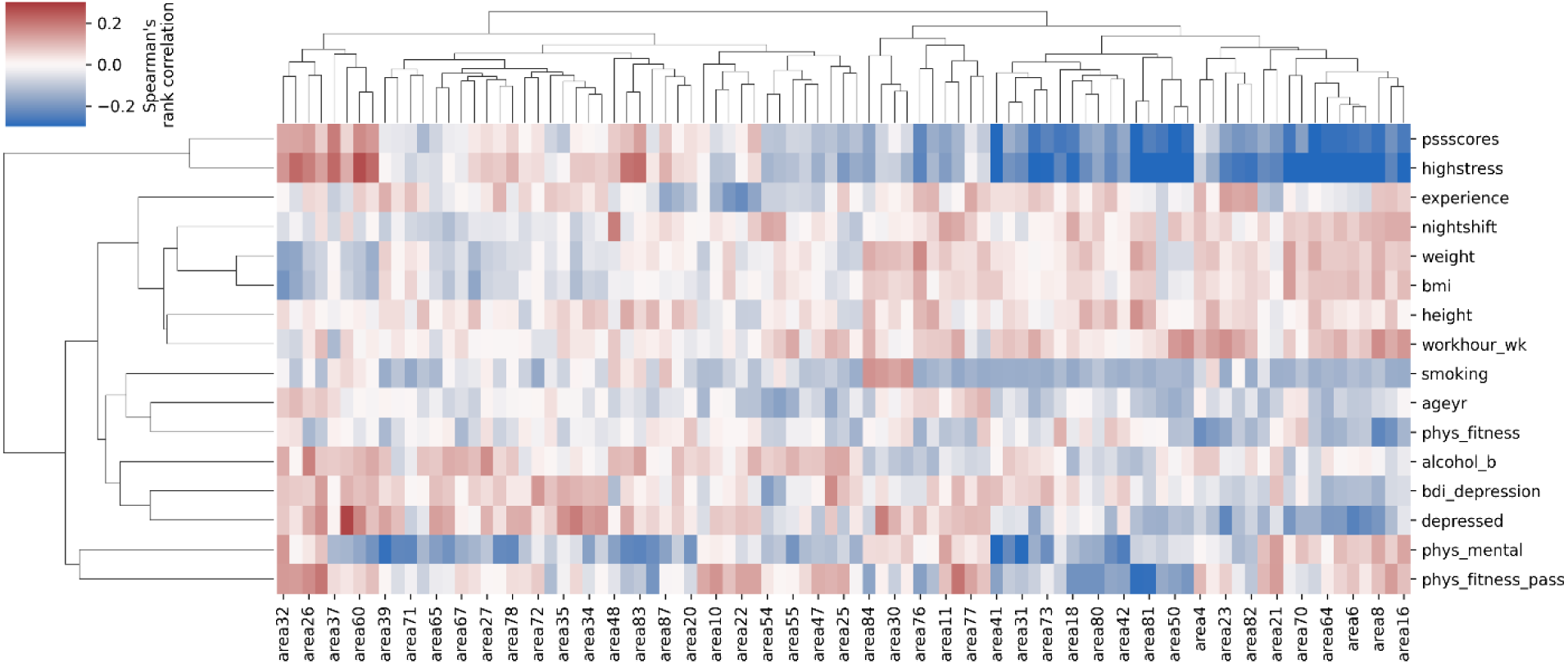
Spearman rank correlation between each VOC and clinical variable. Correlation coefficients were capped at ±0.3 to enhance the contrast. Variables were arranged using hierarchical clustering with Euclidean distance and average linkage.

A multivariable analysis of the association between VOC and stress and depression, via logistic regression and random forest model fitting, indicated that both outcomes can be moderately captured with the best area under the precision-recall curve (AUPRC) of 0.8004 for stress and 0.6532 for depression. This allows us to further analyze and interpret the contribution of each VOC variable on the outcomes. In good agreement with univariate analysis, the best predictor for depression, which is a logistic regression model, utilized only *area88* and *area44* as inputs, both with positive coefficients. Furthermore, area6, area7, and area9 are among the most important variables for the best predictor for stress (**Figure 4**), which is a random forest model. All three VOC variables contribute to predicting the lack of stress as expected. Feature importances from other models are provided in **Supplementary Figure 2**.

**Figure 4.**
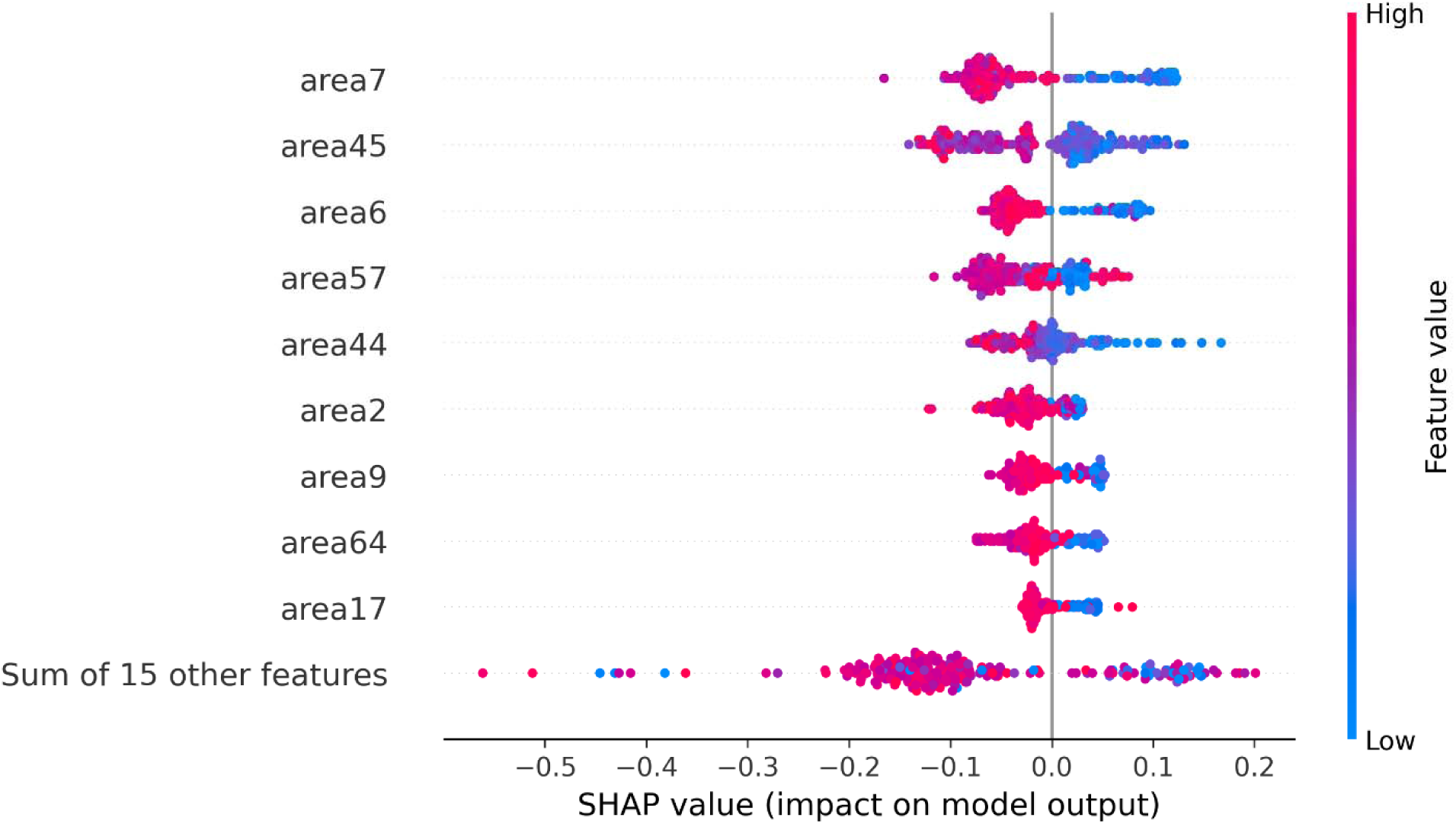
Shapley Additive Explanations (SHAP) values for the best random forest model for predicting stress. Each dot represents a participant. SHAP value indicates whether the prediction leans toward stress (positive value) or no stress (negative value). Color gradient indicates the relative level of VOC among all participants. Variables were ranked by their total absolute SHAP values.

### Dimensional reduction and cluster analysis of VOC

To visualize and interpret the distribution structure of VOC profiles among participants, dimensionality reduction techniques, specifically Principal Component Analysis (PCA) and Uniform Manifold Approximation and Projection (UMAP), were performed. This reveals a clear two-cluster structure (n = 178 and 46) on the first and second components of the PCA that wa later confirmed by hierarchical clustering (**Figure 5** **and Supplementary Figure 3**). Three participants with missing outcome data were excluded from the analysis. One cluster (cluster 2) exhibits a much higher variance on the PCA coordinates and is significantly enriched with participants with diagnosed stress and depression (**Figure 5**, **Table 4**, and **Supplementary Figure 4**, Chi-squared p-values < 3e-5). Among 46 participants in cluster 2, 34 (74%) were diagnosed with high stress and 9 (20%) were diagnosed with depression. In contrast, only 16% and 3% of participants in cluster 1 were diagnosed with high stress and depression, respectively (**Table 4**). There were 55 VOC that were significantly differential between the two clusters, with *area6* and *area9* being the best markers for cluster 1 (**Table 4 and Supplementary Figure 5**). Interestingly, there is no VOC marker for cluster 2 (**Supplementary Figure 6**).

**Figure 5.**
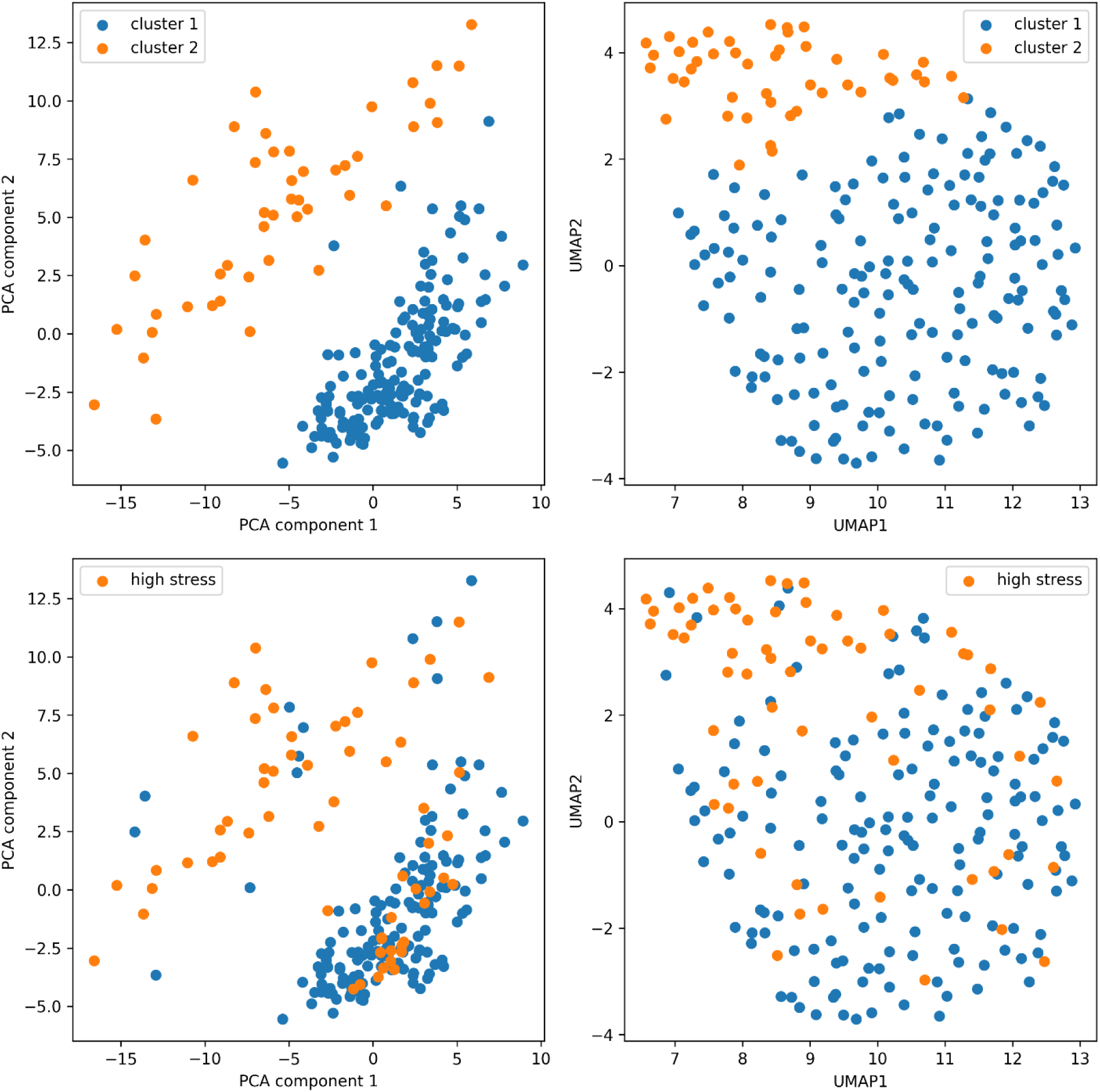
2D scatter plots showing the distribution of VOC profiles on PCA and UMAP coordinates. The two clusters identified by hierarchical clustering and participants with diagnosed stress were indicated.

**Table 4.**
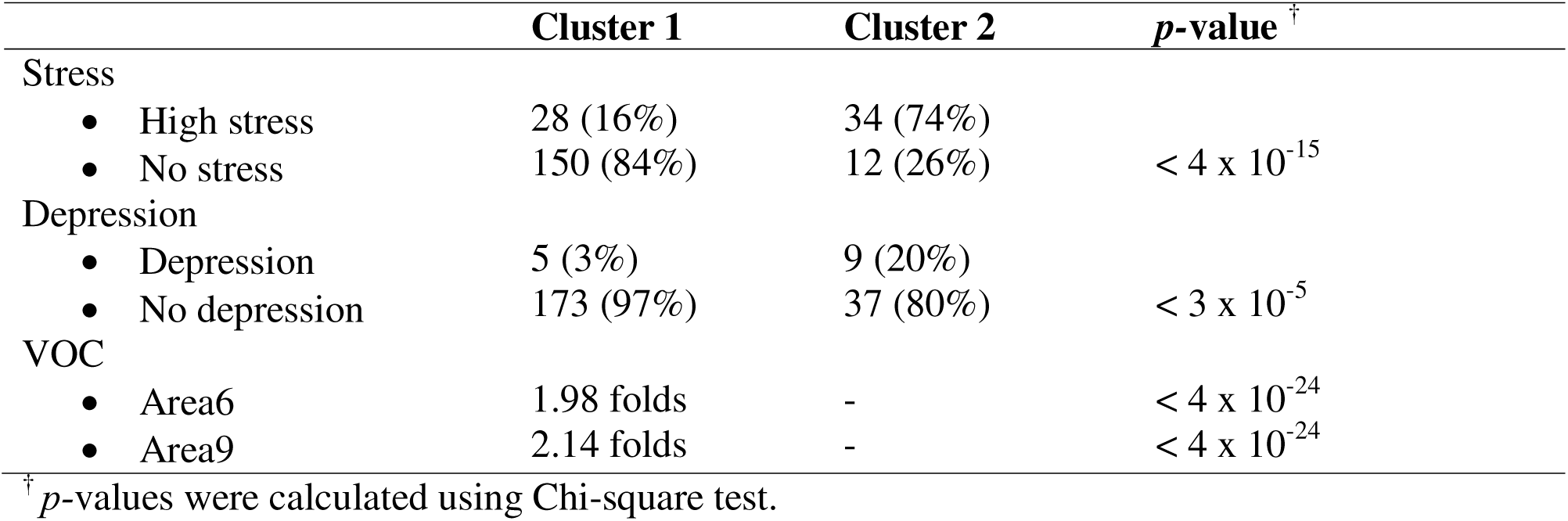
Characteristics of the two VOC clusters.

Next, we investigated whether participants with and without diagnosed high stress from the two clusters exhibited different clinical profiles. This revealed slightly elevated PSS scores among 34 high-stress participants in cluster 2 compared to 28 high-stress participants in cluster 1 (**Supplementary Figure 7**, raw Mann-Whitney U *p*-value = 0.0218). Most interestingly, no-stress participants in cluster 2 also significantly differ from their counterparts in cluster 1. These 12 participants exhibited lower PSS scores (*p*-value = 0.0115) but much higher depression indicator (**Figure 6**, *p*-value < 6 x 10^-6^). All nine participants with diagnosed depression in cluster 2 are among the no-stress group.

**Figure 6.**
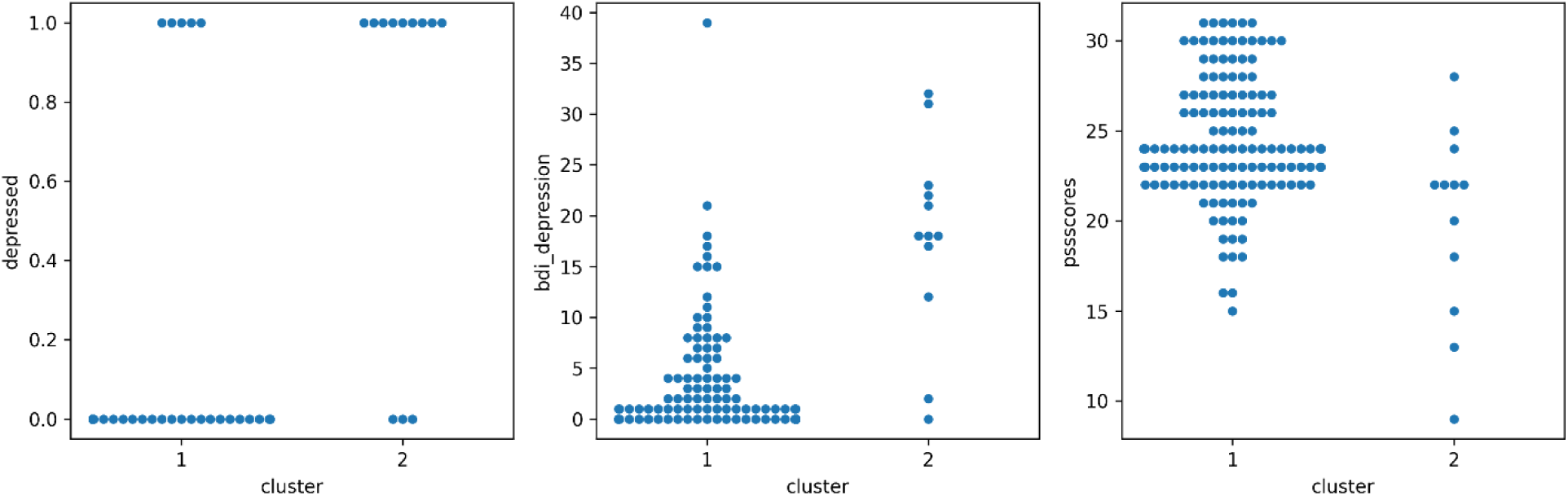
Swarm plots comparing stress and depression indicators between no-stress participants in cluster 1 (n = 150) and cluster 2 (n = 12). All three clinical variables shown significantly differ across clusters (adjusted Mann-Whitney U *p*-value < 0.05).

### Correlation of VOC with microbiota

Possible identities of the proposed markers were examined by injection of authentic standards with their identities listed in **Table 5**. They could be produced according to human or microbiome metabolism/catabolism. For stress markers, it was observed that acetonitrile, octane, pentane and diethyl either showed the same *t*_R_ and *t*_dr_ as that of area 2, areas {7, 9, 57 and 64}, area 17 and area 44, respectively, which leads to the possibility that these four compounds are possible identities of the markers. Based on the available set of standard compounds, only octane and diethyl ether were tentatively identified as potential markers for depression.

**Table 5.**
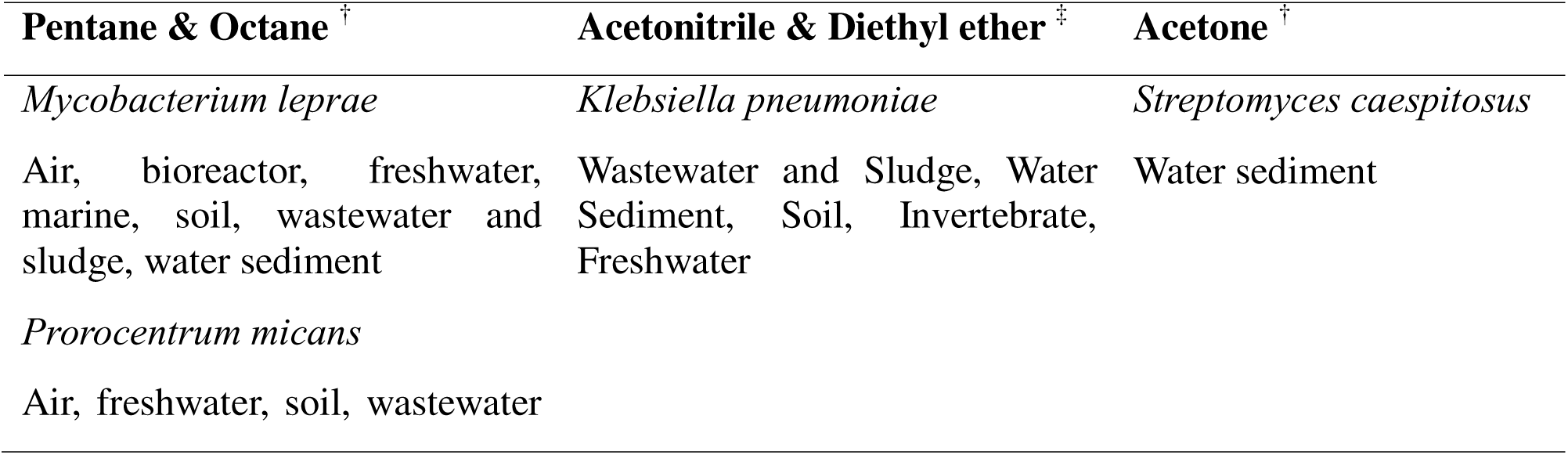
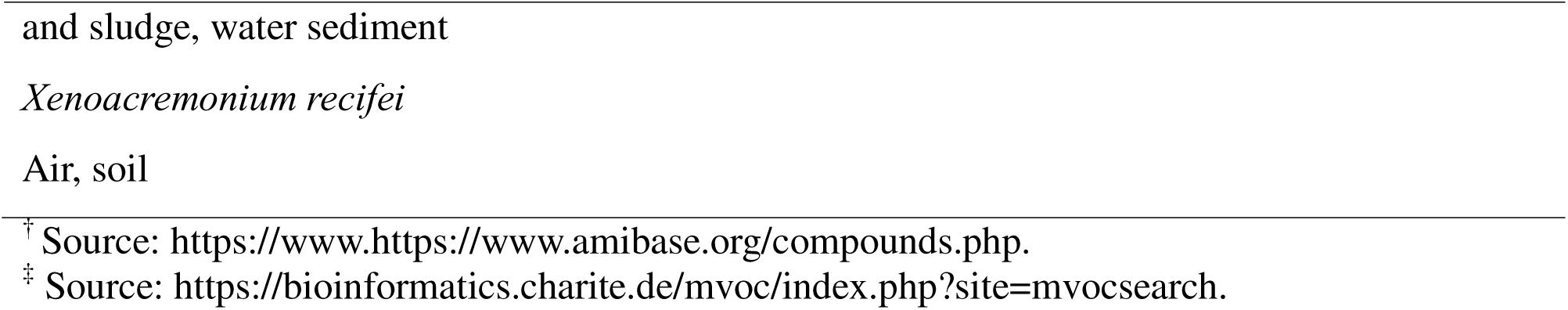
Possible microbiota producing the volatile compound markers.

### Correlation of VOC with Human metabolomic pathway

From the set of the individual marker peaks, the identifiable metabolite set was {pentane, octane, acetonitrile, diethyl ether, and acetone} for high stress condition with the higher to the lower ranks ordered from left to right, respectively. The corresponding set for depression was {diethyl ether, acetonitrile, acetone and octane}. According to the Enrichment analysis using all the database of metabolic pathways freely available in Metaboanalyst and the literature search, the result is documented in **Table 6**.

**Table 6.**
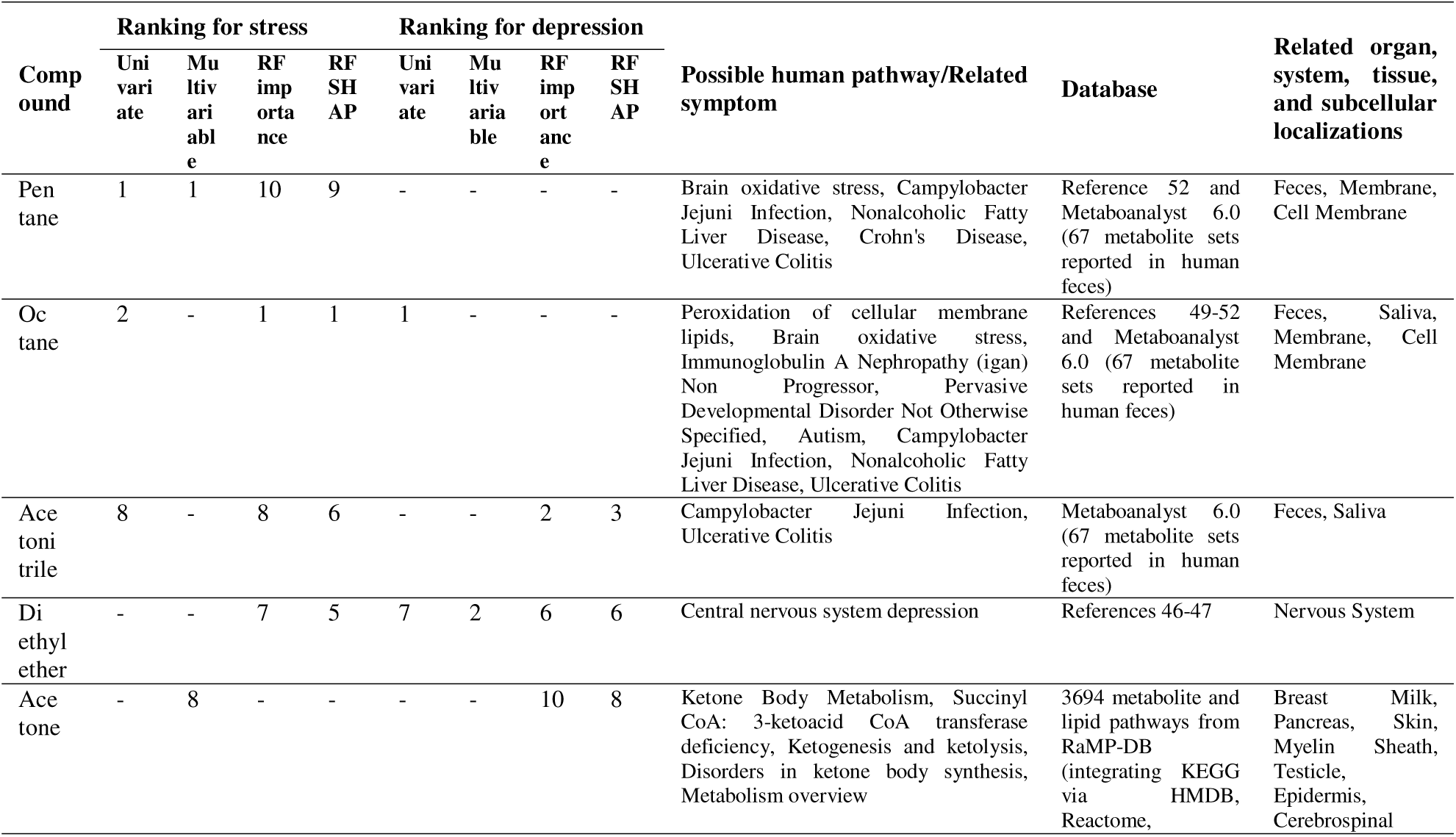
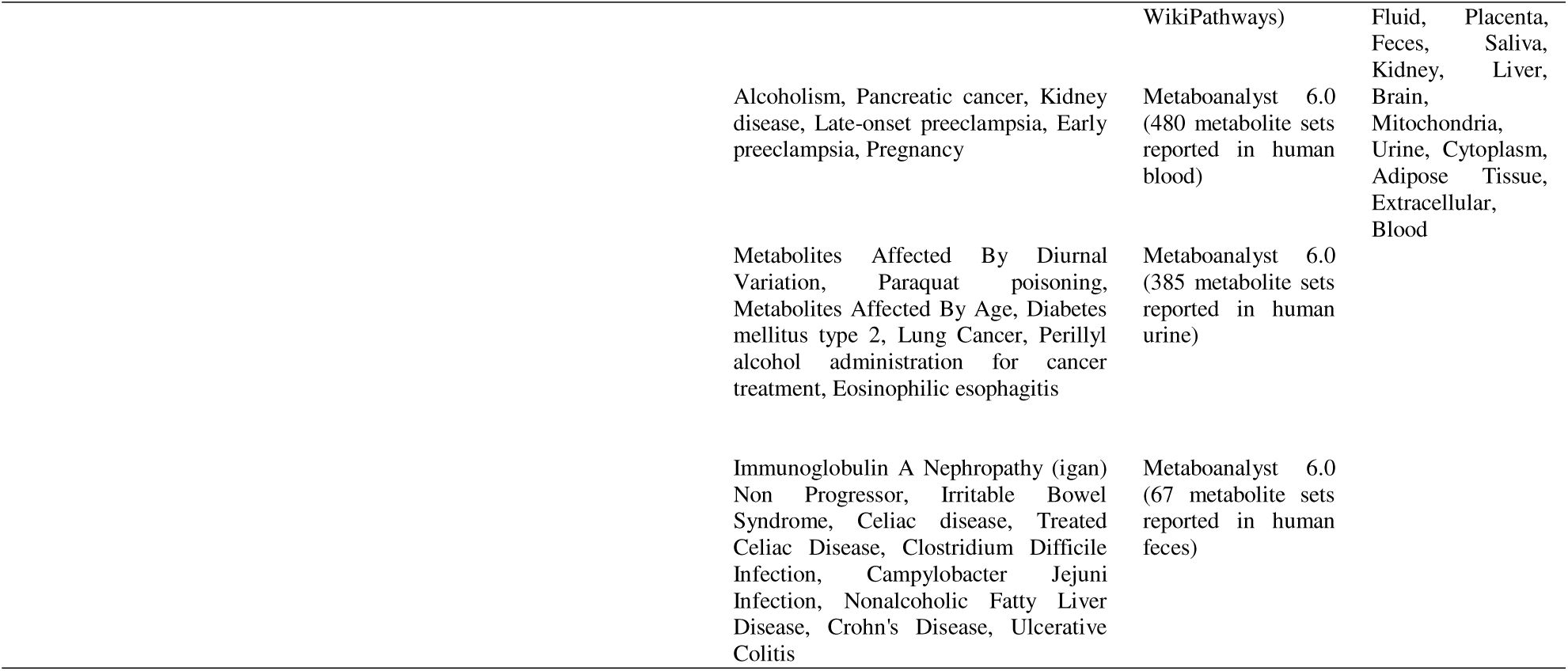
Marker compounds, statistical analysis ranking and possible human metabolic pathways obtained from different references.

## Discussion

In this study, we developed a noninvasive screening method for high stress and depression by analyzing VOCs in axillary sweat. Among 227 firefighters, participants were stratified into high-stress, depression, or normal groups based on validated self-reported questionnaires and psychiatrist-confirmed diagnoses. Using GC-IMS, we identified three VOCs—acetonitrile, octane, and pentane—whose combined levels differentiated high-stress and depression participants from controls with accuracies of 81.3% and 82.8%, respectively. These biomarkers appear to originate from multiple metabolic pathways and potentially involve microbiome-related processes. Importantly, this sweat-based approach demonstrates promise as a rapid, scalable, and objective tool for large-scale mental health screening—particularly in high-risk occupational groups such as firefighters.

### Relationship through human metabolomic pathways

Volatile compounds in human sweat may be related to several metabolic pathways in eccrine, sebaceous or apocrine glands. The skin volatile compounds can be originated from apocrine glands or alternatively obtained from the interactions between skin microorganisms and non-volatile metabolites from the other glands.^23, 42, 43^ Sweat volatiles of high stress and depression participants are expected to be different compared with the negative groups in Bangkok. While octane and pentane were found as common markers found in both fire fighter and nurse population reported in our previous study,^46^ acetonitrile was found as an additional stress marker in sweat of the fire fighters. However, ammonia and methanol in nurses’ sweat were not identified as markers for the fire fighters. This is probably due to different data analysis approaches as well as different genders, locations, sizes and groups of the populations.

Diethyl ether in human blood could be linked to central nervous system depression. Over 90% of diethyl ether is directly eliminated through the lungs, with a smaller amount remaining in sweat. This compound can also metabolize into acetaldehyde and carbon dioxide.^47^ Octane could be potentially linked to oxidative stress involving the peroxidation of cellular membrane lipids ^48^ such as the oxidation of oleic acid producing octanal, nonanal, heptane, and octane.^49–51^ Since chronic stress may contribute to oxidative stress in the brain, ^52^ changes of contents of octane and pentane were thus observed in our study. It should be underscored that major depression and psychological stress are accompanied by increased oxidative stress including lipid peroxidation and that these processes are thought to take part in the pathophysiology of MDD. ^53^

Due to its association with oxidative stress, there have also been attempts to use octane as a biomarker for acute respiratory distress syndrome (ARDS) and pulmonary oxidative injury, but the results showed insufficient diagnostic accuracy. ^54,55^

It should be noted that acetonitrile is not typically produced in the human body under normal conditions. It is introduced through external sources, can be absorbed by all routes, and is rapidly distributed throughout the body. It is then metabolized to cyanide, conjugated with thiosulfate, and eliminated through urine without significant accumulation.^56^ While nitriles, including acetonitrile, can theoretically be formed through oxidation reactions,^57^ there is limited evidence to suggest that this occurs in the human body associated with depression or oxidative stress-related reactions. Other related human metabolomic pathways are listed in **Table 6**.

### Relationship with the skin microbiota

Alternatively, the marker compounds above could also be generated by several microbiota as proposed in **Table 6**. Indeed, pentane and octane could be found along with the local microbiomes (*Mycobacterium leprae* and *Prorocentrum micans*) inside sweat. *K. pneumoniae* could produce acetonitrile and diethyl ether. In addition, nitrile-degrading bacteria, such as *Pseudomonas putida*, could also produce acetonitrile which involves enzymes such as nitrile hydratases and amidases.^58,59^ Major depression is accompanied by increased IgA/IgM responses to *K. pneumoniae* and *K. putida* ^60^ Detection of acetone could possibly indicate presence of *Streptomyces caespitosus* in sweat which could also be observed in water sediment environment.

This study underscores the clinical potential of sweat VOC analysis as a noninvasive method for assessing mental stress and depression. Moreover, exploring the relationship between VOCs and inflammatory markers, such as chemokines and cytokines, may offer deeper insights into the physiological mechanisms linking stress and mental illness. Taken together, these findings hold promise for advancing the diagnostic and prognostic utility of VOC-based approaches in clinical psychiatry.

However, several limitations must be acknowledged. First, the sample consisted exclusively of professional firefighters, who were predominantly male, thus limiting the generalizability of the results to other populations or to female participants. Second, the study’s location in Bangkok may introduce potential confounders related to regional lifestyle and environmental exposures that could influence VOC profiles. Third, although smokers were not initially excluded, all participants in the final sample were non-smokers, potentially narrowing the range of detectable VOCs and limiting the applicability of the results to populations with diverse smoking behaviors. Finally, the sensitivity of GC-IMS in detecting certain biomarkers at very low concentrations poses challenges in capturing subtle variations in complex biological samples. Future research should explore the influence of diverse geographical regions and environmental factors on VOC profiles to enhance generalizability.

## Conclusion

Sweat based stress and depression screening approach using GC-IMS was demonstrated with >80% accuracy. This relies on detection of different volatile marker peaks in headspaces of axillary sweat samples obtained from firefighters. By injection of authentic standards, the possible identities of these highly volatile components were obtained which could be the metabolites originated from several metabolic pathways in humans as well as commensal microorganisms found in the human body.

## Data Availability

All data produced in the present study are available upon reasonable request to the authors.

## Acknowledgement

This study was funded by the Medical Council of Thailand (Grant No. 012/1621). This research project is supported by The Second Century Fund (C2F), Chulalongkorn University and Maha Chakri Sirindhorn Clinical Research Center Under the Royal Patronage, Faculty of Medicine, Chulalongkorn University, Bangkok, Thailand. Sci Spec Company Limited, Bangkok, Thailand provided the GC–IMS instrument. The authors also thank all firefighters from the Bangkok fire and rescue department, the Bangkok Metropolitan Administration (BMA) who volunteered to participate in this study; the staff members that helped to recruit participants and data collection, in particular, Mrs. Jenjira Suwongsa, Mrs. Chunyanuch Boonsuwan, Mr. Pongpipat Nopakaow, and Mrs. Friscilla Hermatasia.

## REFERENCES

1. Carey MG, Al-Zaiti SS, Dean GE, Sessanna L, Finnell DS. Sleep problems, depression, substance use, social bonding, and quality of life in professional firefighters. J Occup Environ Med. 2011;53:928–33.

2. Wolffe TAM, Robinson A, Clinton A, Turrell L, Stec AA. Mental health of UK firefighters. Scientific Reports. 2023;13:62.

3. Harvey SB, Milligan-Saville JS, Paterson HM, Harkness EL, Marsh AM, Dobson M, et al. The mental health of fire-fighters: An examination of the impact of repeated trauma exposure. Aust N Z J Psychiatry. 2016;50:649–58.

4. Stanley IH, Hom MA, Hagan CR, Joiner TE. Career prevalence and correlates of suicidal thoughts and behaviors among firefighters. J Affect Disord. 2015;187:163–71.

5. Park H, Kim JI, Min B, Oh S, Kim J-H. Prevalence and correlates of suicidal ideation in Korean firefighters: a nationwide study. BMC Psychiatry. 2019;19:428.

6. Stanley IH, Boffa JW, Smith LJ, Tran JK, Schmidt NB, Joiner TE, et al. Occupational stress and suicidality among firefighters: Examining the buffering role of distress tolerance. Psychiatry Research. 2018;266:90–6.

7. Carleton RN, Afifi TO, Turner S, Taillieu T, Duranceau S, LeBouthillier DM, et al. Mental Disorder Symptoms among Public Safety Personnel in Canada. Can J Psychiatry. 2018;63:54– 64.

8. Chen X, Zhang L, Peng Z, Chen S. Factors Influencing the Mental Health of Firefighters in Shantou City, China. Psychology Research and Behavior Management. 2020;13:529–36.

9. Wagner SL, White N, Buys N, Carey MG, Corneil W, Fyfe T, et al. Systematic review of mental health symptoms in firefighters exposed to routine duty-related critical incidents. Traumatology. 2021;27:285–302.

10. Di Nota PM, Kasurak E, Bahji A, Groll D, Anderson GS. Coping among public safety personnel: A systematic review and meta–analysis. Stress and Health. 2021;37:613–30.

11. Pennington ML, Carpenter TP, Synett SJ, Torres VA, Teague J, Morissette SB, et al. The Influence of Exposure to Natural Disasters on Depression and PTSD Symptoms among Firefighters. Prehosp Disaster Med. 2018;33:102–8.

12. Ricciardelli R, Czarnuch S, Afifi TO, Taillieu T, Carleton RN. Public Safety Personnel’s interpretations of potentially traumatic events. Occup Med (Lond). 2020;70:155–61.

13. Cramm H, Richmond R, Jamshidi L, Edgelow M, Groll D, Ricciardelli R, et al. Mental Health of Canadian Firefighters: The Impact of Sleep. Int J Environ Res Public Health. 2021;18.

14. Herzog J, O’Dare K, King E, Rotunda R, Dillard D. Trauma symptoms and suicidal ideation in firefighters with consideration of exposure to natural disaster. Traumatology. 2022:No Pagination Specified–No Pagination Specified.

15. Saijo Y, Ueno T, Hashimoto Y. Job stress and depressive symptoms among Japanese fire fighters. Am J Ind Med. 2007;50:470–80.

16. De Pessemier B, Grine L, Debaere M, Maes A, Paetzold B, Callewaert C. Gut-Skin Axis: Current Knowledge of the Interrelationship between Microbial Dysbiosis and Skin Conditions. Microorganisms. 2021;9.

17. Arck P, Handjiski B, Hagen E, Pincus M, Bruenahl C, Bienenstock J, et al. Is there a ’gut-brain-skin axis’? Exp Dermatol. 2010;19:401–5.

18. Arck P, Paus R. From the brain-skin connection: the neuroendocrine-immune misalliance of stress and itch. Neuroimmunomodulation. 2006;13:347–56.

19. Hacimusalar Y, Eşel E. Suggested Biomarkers for Major Depressive Disorder. Noro Psikiyatr Ars. 2018;55:280–90.

20. Dhama K, Latheef SK, Dadar M, Samad HA, Munjal A, Khandia R, et al. Biomarkers in Stress Related Diseases/Disorders: Diagnostic, Prognostic, and Therapeutic Values. Front Mol Biosci. 2019;6:91.

21. Lee S, Kim H, Park MJ, Jeon HJ. Current Advances in Wearable Devices and Their Sensors in Patients With Depression. Frontiers in Psychiatry. 2021;12.

22. Fathizadeh H, Taghizadeh S, Safari R, Khiabani SS, Babak B, Hamzavi F, et al. Study presence of COVID-19 (SARS-CoV-2) in the sweat of patients infected with Covid-19. Microbial Pathogenesis. 2020;149:104556.

23. Wysocki CJ, Preti G. Facts, fallacies, fears, and frustrations with human pheromones. *The Anatomical Record Part A: Discoveries in Molecular*, Cellular, and Evolutionary Biology. 2004;281A:1201–11.

24. Bernier UR, Kline DL, Barnard DR, Schreck CE, Yost RA. Analysis of Human Skin Emanations by Gas Chromatography/Mass Spectrometry. 2. Identification of Volatile Compounds That Are Candidate Attractants for the Yellow Fever Mosquito (Aedes aegypti). Analytical Chemistry. 2000;72:747–56.

25. Davis C, Pleil J, Beauchamp J. Breathborne Biomarkers and the Human Volatilome: Elsevier Science; 2020.

26. Gallagher M, Wysocki CJ, Leyden JJ, Spielman AI, Sun X, Preti G. Analyses of volatile organic compounds from human skin. The British journal of dermatology. 2008;159:780–91.

27. Verhulst NO, Andriessen R, Groenhagen U, Bukovinszkiné Kiss G, Schulz S, Takken W, et al. Differential Attraction of Malaria Mosquitoes to Volatile Blends Produced by Human Skin Bacteria. PLOS ONE. 2011;5:e15829.

28. Amibase Anmicro. Natural compounds. [Internet]. [cited 2024 Aug 16] Available from : https://www.amibase.org/compounds.php.

29. Martin HJ, Turner MA, Bandelow S, Edwards L, Riazanskaia S, Thomas CLP. Volatile organic compound markers of psychological stress in skin: a pilot study. Journal of Breath Research. 2016;10:046012.

30. Zamkah A, Hui T, Andrews S, Dey N, Shi F, Sherratt RS. Identification of Suitable Biomarkers for Stress and Emotion Detection for Future Personal Affective Wearable Sensors. Biosensors. 2020;10:40.

31. Herrero M, Ibáñez E, Cifuentes A, Bernal J. Multidimensional chromatography in food analysis. Journal of Chromatography A. 2009;1216:7110–29.

32. Cannon RJ, Agyemang D, Curto NL, Yusuf A, Chen MZ, Janczuk AJ. In-depth analysis of Ciflorette strawberries (Fragaria × ananassa ‘Ciflorette’) by multidimensional gas chromatography and gas chromatography-olfactometry. Flavour and Fragrance Journal. 2015;30:302–19.

33. Ruszkiewicz DM, Sanders D, O’Brien R, Hempel F, Reed MJ, Riepe AC, et al. Diagnosis of COVID-19 by analysis of breath with gas chromatography-ion mobility spectrometry - a feasibility study. EClinicalMedicine. 2020;29–30:100609.

34. Dorota M. Ruszkiewicz, Austin Meister, Renelle Myers, Ion Mobility Spectrometry in Clinical and Emergency Setting: Research and Potential Applications, in: Stefan Weigl (Eds.), Breath Analysis: An Approach for Smart Diagnostics, Springer, Cham, 2022, pp. 45–71.

35. Vautz W, Seifert L, Mohammadi M, et al. Detection of axillary perspiration metabolites using ion mobility spectrometry coupled to rapid gas chromatography. Anal Bioanal Chem. 2020;412:223–32.

36. Wongpakaran N, Wongpakaran T. The Thai version of the PSS-10: An Investigation of its psychometric properties. BioPsychoSocial Medicine. 2010;4:6.

37. Lotrakul M, Sukanich P. Development of the Thai Depression Inventory. J Med Assoc Thai. 1999;82:1200–7.

38. Stauder A, Thege BK, editors. SUMMARY ON THE VALIDITY STUDY OF THE HUNGARIAN VERSION OF THE PERCEIVED STRESS SCALE 2007.

39. Lee E-H. Review of the Psychometric Evidence of the Perceived Stress Scale. Asian Nursing Research. 2012;6:121–7.

40. Wang YP, Gorenstein C. Psychometric properties of the Beck Depression Inventory-II: a comprehensive review. Braz J Psychiatry. 2013;35:416–31.

41. Cohen S, Kamarck T, Mermelstein R. A Global Measure of Perceived Stress. Journal of Health and Social Behavior. 1983;24:385–96.

42. Hanioka T, Ojima M, Tanaka K, Aoyama H. Relationship between smoking status and tooth loss: Findings from national databases in Japan. Journal of Epidemiology. 2007;17:125–32.

43. Brinkman GL, Coates EO, Jr. The effect of bronchitis, smoking, and occupation on ventilation. Am Rev Respir Dis. 1963;87:684–93.

44. Labows JN, Mcginley KJ, Kligman AM. Perspectives on axillary odor. J Soc Cosmet Chem. 1982;33:193–202.

45. Wackett LP. Microbial biocatalysis databases. Microbial Biotechnology. 2018;11:429–31.

46. Tungkijanansin N, Sirinara P, Tunvirachaisakul C, Srikam S, Kittiban K, Thongthip S, et al. Sweat-based stress screening with gas chromatography-ion mobility spectrometry and electronic nose. Anal Chim Acta. 2024;1320:343029.

47. Cox D, DeRienz R, Jufer Phipps RA, Levine B, Jacobs A, Fowler D. Distribution of Ether in Two Postmortem Cases. Journal of Analytical Toxicology. 2006;30:635–7.

48. Tranchito L, Albert CL, Gul Z, Cikach F, Grove D, Dweik R, et al. Exhaled Acetone and Pentane in Patients Undergoing Cardiopulmonary Exercise Testing. Journal of Cardiac Failure. 2017;23:S31.

49. Wisthaler A, Weschler CJ. Reactions of ozone with human skin lipids: Sources of carbonyls, dicarbonyls, and hydroxycarbonyls in indoor air. Proceedings of the National Academy of Sciences. 2010;107:6568–75.

50. Mochalski P, Unterkofler K, Teschl G, Amann A. Potential of volatile organic compounds as markers of entrapped humans for use in urban search-and-rescue operations. TrAC Trends in Analytical Chemistry. 2015;68:88–106.

51. Kneepkens CM, Lepage G, Roy CC. The potential of the hydrocarbon breath test as a measure of lipid peroxidation. Free Radic Biol Med. 1994;17:127–60.

52. Juszczyk G, Mikulska J, Kasperek K, Pietrzak D, Mrozek W, Herbet M. Chronic Stress and Oxidative Stress as Common Factors of the Pathogenesis of Depression and Alzheimer’s Disease: The Role of Antioxidants in Prevention and Treatment. Antioxidants. 2021;10:1439.

53. Maes M, Galecki P, Chang YS, Berk M. A review on the oxidative and nitrosative stress (O&NS) pathways in major depression and their possible contribution to the (neuro)degenerative processes in that illness. Prog Neuropsychopharmacol Biol Psychiatry. 2011 Apr 29;35(3):676–92. doi: 10.1016/j.pnpbp.2010.05.004. Epub 2010 May 12. PMID: 20471444.

54. Hagens LA, Heijnen NFL, Smit MR, Verschueren ARM, Nijsen TME, Geven I, et al. Octane in exhaled breath to diagnose acute respiratory distress syndrome in invasively ventilated intensive care unit patients. ERJ Open Research. 2023;9:00214–2023.

55. Fenn D, Lilien TA, Hagens LA, Smit MR, Heijnen NFL, Tuip-de Boer AM, et al. Validation of volatile metabolites of pulmonary oxidative injury: a bench to bedside study. ERJ Open Research. 2022:00427–2022.

56. Robles H. Acetonitrile. In: Wexler P, editor. Encyclopedia of Toxicology (Fourth Edition). Oxford: Academic Press; 2024. p. 79–82.

57. Bellale EV, Huddar SN, Mahajan US, Akamanchi KG. Oxidative decarboxylation of α-amino acids to nitriles using o-iodoxybenzoic acid in aqueous ammonia. 2011;83:607–12.

58. Nawaz MS, Chapatwala KD, Wolfram JH. Degradation of Acetonitrile by Pseudomonas putida. Applied and environmental microbiology. 1989;55:2267–74.

59. Heidari A, Asoodeh A. A novel nitrile-degrading enzyme (nitrile hydratase) from Ralstonia sp.ZA96 isolated from oil-contaminated soils. Biocatalysis and Agricultural Biotechnology. 2019;21:101285.

60. Maes M, Kubera M, Leunis JC. The gut-brain barrier in major depression: intestinal mucosal dysfunction with an increased translocation of LPS from gram-negative enterobacteria (leaky gut) plays a role in the inflammatory pathophysiology of depression. Neuro Endocrinol Lett. 2008;29(1):117–24.

59. Peyrovian B, Rosenblat JD, Pan Z, Iacobucci M, Brietzke E, McIntyre RS. The glycine site of NMDA receptors: A target for cognitive enhancement in psychiatric disorders. Prog Neuropsychopharmacol Biol Psychiatry 2019;92:387–404.

60. Hashimoto K, Yoshida T, Ishikawa M, Fujita Y, Niitsu T, Nakazato M, et al. Increased serum levels of serine enantiomers in patients with depression. Acta Neuropsychiatr. 2016;28:173–8.

61. Altamura C, Maes M, Dai J, Meltzer HY. Plasma concentrations of excitatory amino acids, serine, glycine, taurine and histidine in major depression. Eur Neuropsychopharmacol. 1995;5 Suppl:71-5.

62. Yamamori H, Hashimoto R, Fujita Y, Numata S, Yasuda Y, Fujimoto M, et al. Changes in plasma D-serine, L-serine, and glycine levels in treatment-resistant schizophrenia before and after clozapine treatment. Neurosci Lett. 2014;582:93–8.

63. Moradi F, Lotfi K, Armin M, Clark CCT, Askari G, Rouhani MH. The association between serum homocysteine and depression: A systematic review and meta-analysis of observational studies. Eur J Clin Invest. 2021;51:e13486.

64. Bhatia P, Singh N. Homocysteine excess: delineating the possible mechanism of neurotoxicity and depression. Fundam Clin Pharmacol. 2015;29:522–8.

65. Sawai A, Ohshige K, Kura N, Tochikubo O. Influence of mental stress on the plasma homocysteine level and blood pressure change in young men. Clin Exp Hypertens. 2008;30:233–41.

66. Stoney CM. Plasma homocysteine levels increase in women during psychological stress. Life Sci. 1999;64:2359–65.

67. Kuebler U, Linnebank M, Semmler A, Stoffel-Wagner B, La Marca R, Ehlert U, et al. Plasma homocysteine levels increase following stress in older but not younger men. Psychoneuroendocrinology. 2013;38:1381–7.

68. Crombez EA, Cederbaum SD. Hyperargininemia due to liver arginase deficiency. Mol Genet Metab. 2005;84:243–51.

69. Trueb L, Lepori M, Duplain H, Scherrer U, Sartori C. Nitric oxide mediates the blood pressure response to mental stress in humans. Swiss Med Wkly. 2012;142:w13627.

70. Bélanger-Quintana A, Arrieta Blanco F, Barrio-Carreras D, Bergua Martínez A, Cañedo Villarroya E, García-Silva MT, et al. Recommendations for the Diagnosis and Therapeutic Management of Hyperammonaemia in Paediatric and Adult Patients. Nutrients. 2022;14.

71. Duan Y, Wu X, Liang S, Jin F. Elevated Blood Ammonia Level Is a Potential Biological Risk Factor of Behavioral Disorders in Prisoners. Behav Neurol. 2015;2015:1–5.

